# Development and validation of a clinical and genetic model for predicting risk of severe COVID-19

**DOI:** 10.1101/2021.03.09.21253237

**Authors:** Gillian S. Dite, Nicholas M. Murphy, Richard Allman

## Abstract

Clinical and genetic risk factors for severe COVID-19 are often considered independently and without knowledge of the magnitudes of their effects on risk. Using SARS-CoV-2 positive participants from the UK Biobank, we developed and validated a clinical and genetic model to predict risk of severe COVID-19. We used multivariable logistic regression on a 70% training dataset and used the remaining 30% for validation. We also validated a previously published prototype model. In the validation dataset, our new model was associated with severe COVID-19 (odds ratio per quintile of risk=1.77, 95% confidence interval [CI]=1.64, 1.90) and had excellent discrimination (area under the receiver operating characteristic curve=0.732, 95% CI=0.708, 0.756). We assessed calibration using logistic regression of the log odds of the risk score, and the new model showed no evidence of over- or under-estimation of risk (α=−0.08; 95% CI=−0.21, 0.05) and no evidence or over- or under-dispersion of risk (β=0.90, 95% CI=0.80, 1.00). Accurate prediction of individual risk is possible and will be important in regions where vaccines are not widely available or where people refuse or are disqualified from vaccination, especially given uncertainty about the extent of infection transmission among vaccinated people and the emergence of SARS-CoV-2 variants of concern.

**Key results:**

- Accurate prediction of the risk of severe COVID-19 can inform public heath interventions and empower individuals to make informed choices about their day-to-day activities.
- Age and sex alone do not accurately predict risk of severe COVID-19.
- Our clinical and genetic model to predict risk of severe COVID-19 performs extremely well in terms of discrimination and calibration.

## Introduction

The COVID-19 pandemic continues to dominate global public health, with countries having varying success with infection control measures and social distancing protocols [1], Coupled with this are the logistical challenges with the distribution of vaccines [2] and the emergence of SARS-CoV-2 variants of concern [3, 4]. Of those who become infected with SARS-CoV-2, 10–15% will develop severe COVID-19 requiring hospitalisation and 5% will require intensive care [5]. At all stages of the pandemic, there has been an urgent need for accurate quantification of risk of severe COVID-19 to inform protection from infection for those at increased risk.

Epidemiological analyses have recognized that sex and increasing age are risk factors for severe COVID-19 and that common medical comorbidities contribute to individual risk [6-8]. Our previous analysis showed that the effects of sex and age are attenuated when comorbidities are taken into account [9]. The effect of human genetic variation on COVID-19 severity has been examined by the COVID-19 Host Genetics Initiative, which has now released several meta-analyses of the available genome-wide association studies of COVID-19 severity [10, 11]. Using population controls, Ellinghaus et al. [12] identified two loci (3p21.31 and 9q34.2) as being strongly associated with respiratory failure from COVID-19 and Shelton et al. [13] identified the 3p21.31 locus as being associated with severe COVID-19. Also using population controls, Pairo-Castineira et al. [14] identified eight single-nucleotide polymorphisms (SNPs) that achieved genome-wide significance for intensive care admission and identified six SNPs (two of which were also in the panel of eight SNPs) associated with risk of hospitalization.

The emergency authorization of SARS-CoV-2 vaccines [15] does not diminish the value of accurate prediction of individual risk of severe COVID-19. Extensive vaccine disqualification criteria (such as pre-existing conditions, pregnancy, and age), vaccine hesitancy, uncertainty as to whether the vaccines are effective against emerging variants of concern [4], and an unknown extent to which vaccines prevent the transmission of infection mean that many people will be at risk of severe COVID-19 should they become infected with SARS-CoV-2.

We previously developed a prototype risk model [9] based upon early data from the UK Biobank [16, 17] and SNPs identified from the COVID-19 Host Genetics Initiative Release 2 meta-analysis of hospitalized vs non-hospitalized COVID-19 cases (which was at that time almost exclusively UK Biobank samples) [10, 18]. Our prototype model appeared to perform well but was based on a small sample size from the first wave of the pandemic [9].

We decided not to attempt validation in this dataset because of our concern about the representativeness of the data (the SARS-CoV-2 testing data was ascertained early in the pandemic when the limitations on testing availability in the United Kingdom meant that mild and asymptomatic cases were not identified).

In the interim, the UK Biobank has released further data from participants confirmed to be infected with SARS-CoV-2. This latest data release (2205 cases and 5416 controls) has a larger proportion of non-hospitalized people, providing more confidence that they are a more representative. In this paper, we perform a validation study of our prototype model and to develop and validate a new clinical and genetic model to predict risk of severe COVID-19.

## Methods

### UK Biobank data and eligibility

Since our first paper on the development of a risk prediction model for severe COVID-19 [9], the UK Biobank [16, 17] has accumulated a large number of additional SARS-CoV-2 test results [19]. For this analysis, we downloaded an updated results file on 8 January 2021. As in our first paper, eligible participants were active UK Biobank participants with a positive SARS-CoV-2 test result and who had SNP and hospital data available [9]. Of the 47990 UK Biobank participants with at least one SARS-CoV-2 test result, 8672 (18.1%) had a positive test result, and of these, 7621 met our eligibility criteria.

As we did previously [9], we used source of test result as a proxy for severity of disease, where inpatient results were considered severe disease (cases) and outpatient results were considered non-severe disease (controls). If a participant had more than one test result, we classified them as having severe disease if at least one of their results was from an inpatient setting. Of the 7621 eligible participants, 2205 (28.9%) were cases and 5416 (71.7%) were controls.

### Data extraction

We used UK Biobank clinical and genetic data that we had previously downloaded (see Table 1). We used Plink version 1.9 [20, 21] to extract SNP data from the UK Biobank imputation dataset. We extracted genotypes of the 64 SNPs that were used to calculate the SNP score in our prototype model [9] and the 12 SNPs from Pairo-Castineira et al. [14] We also identified 43 SNPs from the B1_ALL (hospitalized versus non-hospitalized cases of COVID-19) results of the COVID19-hg GWAS meta-analyses round 4, conducted by the COVID-19 Host Genetics Initiative consortium.[10, 22] These SNPs were selected by pruning variants with a P value of greater than 10^−5^ and linkage disequilibrium variants that had an R^2^ of greater than 0.5 for all populations. Of these 43 SNPS, 40 were available for extraction in the UK Biobank imputation dataset. The SNPs considered in the current paper are listed in Supplementary Table S1.

**Table 1.** Characteristics of cases and controls in the training and validation datasets for the variables considered for inclusion in the new model.

| Variable |  | Training |  | Validation |  |
| --- | --- | --- | --- | --- | --- |
|  |  | Cases<br>N=1,544 | Controls<br>N=3,791 | Cases<br>N=661 | Controls<br>N=1,625 |
|  |  | Mean (SD) | Mean (SD) | Mean (SD) | Mean (SD) |
| Inverse of body mass index | 10/(kg/m<sup>2</sup>) | 0.35 (0.06) | 0.37 (0.06) | 0.35 (0.06) | 0.36 (0.06) |
|  |  | N (%) | N (%) | N (%) | N (%) |
| Age group (years) | 50–54 | 97 (6.3) | 465 (12.3) | 40 (6.1) | 192 (11.8) |
|  | 55–59 | 178 (11.5) | 872 (23.0) | 85 (12.9) | 401 (24.7) |
|  | 60–64 | 144 (9.3) | 668 (17.6) | 70 (10.6) | 290 (17.9) |
|  | 65–69 | 197 (12.8) | 578 (15.3) | 83 (12.6) | 240 (14.8) |
|  | 70–74 | 343 (22.2) | 589 (15.5) | 127 (19.2) | 247 (15.2) |
|  | 75–79 | 436 (28.2) | 481 (12.7) | 190 (28.7) | 196 (12.1) |
|  | 80+ | 149 (9.7) | 138 (3.6) | 66 (10.0) | 59 (3.6) |
| Sex | Female | 665 (43.1) | 2,080 (54.9) | 281 (42.5) | 857 (52.7) |
|  | Male | 879 (56.9) | 1,711 (45.1) | 380 (57.5) | 768 (47.3) |
| Ethnicity | White | 1,381 (89.4) | 3,481 (91.8) | 599 (90.6) | 1,486 (91.5) |
|  | Other/Unknown | 163 (10.6) | 310 (8.2) | 62 (9.4) | 139 (8.6) |
|  |  | Cases<br>N=1,544 | Controls<br>N=3,791 | Cases<br>N=661 | Controls<br>N=1,625 |
| ABO blood type | O | 627 (40.6) | 1,472 (38.8) | 300 (45.4) | 640 (39.4) |
|  | A | 701 (45.4) | 1,764 (46.5) | 265 (40.1) | 750 (46.2) |
|  | B | 164 (10.6) | 393 (10.4) | 70 (10.6) | 169 (10.4) |
|  | AB | 52 (3.4) | 162 (4.3) | 26 (3.9) | 66 (4.1) |
| Asthma | No | 1,286 (83.3) | 3,355 (88.5) | 549 (83.1) | 1,403 (86.3) |
|  | Yes | 258 (16.7) | 436 (11.5) | 112 (16.9) | 222 (13.7) |
| Autoimmune disease (rheumatoid arthritis, lupus or psoriasis) | No | 1,448 (93.8) | 3,654 (96.4) | 616 (93.2) | 1,571 (96.7) |
|  | Yes | 96 (6.2) | 137 (3.6) | 45 (6.8) | 54 (3.3) |
| Cancer – haematological | No | 1,494 (96.8) | 3,765 (99.3) | 637 (96.4) | 1,615 (99.4) |
|  | Yes | 50 (3.2) | 26 (0.7) | 24 (3.6) | 10 (0.6) |
| Cancer – non-haematological | No | 1,217 (78.8) | 3,323 (87.7) | 525 (79.4) | 1,425 (87.7) |
|  | Yes | 327 (21.2) | 468 (12.4) | 136 (20.6) | 200 (12.3) |
| Cerebrovascular disease | No | 1,338 (86.7) | 3,626 (95.7) | 565 (85.5) | 1,555 (95.7) |
|  | Yes | 206 (13.3) | 165 (4.4) | 96 (14.5) | 70 (4.3) |
| Diabetes | No | 1,168 (75.7) | 3,453 (91.1) | 525 (79.4) | 1,470 (90.5) |
|  | Yes | 376 (24.4) | 338 (8.9) | 136 (20.6) | 155 (9.5) |
| Heart disease | No | 1,013 (65.6) | 3,205 (84.5) | 454 (68.7) | 1,374 (84.6) |
|  | Yes | 531 (34.4) | 586 (15.5) | 207 (31.3) | 251 (15.5) |
| Hypertension | No | 679 (44.0) | 2,661 (70.2) | 304 (46.0) | 1,134 (69.8) |
|  | Yes | 865 (56.0) | 1,130 (29.8) | 357 (54.0) | 491 (30.2) |
|  |  | Cases<br>N=1,544 | Controls<br>N=3,791 | Cases<br>N=661 | Controls<br>N=1,625 |
| Immunocompromised | No | 1,525 (98.8) | 3,780 (99.7) | 653 (98.8) | 1,620 (99.7) |
|  | Yes | 19 (1.2) | 11 (0.3) | 8 (1.2) | 5 (0.3) |
| Kidney disease | No | 1,318 (85.4) | 3,677 (97.0) | 581 (87.9) | 1,562 (96.1) |
|  | Yes | 226 (14.6) | 114 (3.0) | 80 (12.1) | 63 (3.9) |
| Liver disease | No | 1,442 (93.4) | 3,683 (97.2) | 613 (92.7) | 1,579 (97.2) |
|  | Yes | 102 (6.6) | 108 (2.9) | 48 (7.3) | 46 (2.8) |
| Respiratory disease (excluding asthma) | No | 1,026 (66.5) | 3,487 (92.0) | 448 (67.8) | 1,489 (91.6) |
|  | Yes | 518 (33.6) | 304 (8.0) | 213 (32.2) | 136 (8.4) |
| rs112641600 | C/C | 1,249 (80.9) | 2,972 (78.4) | 525 (79.4) | 1,271 (78.2) |
|  | T/C | 262 (17.0) | 708 (18.7) | 120 (18.2) | 317 (19.5) |
|  | T/T | 11 (0.7) | 58 (1.5) | 5 (0.76) | 20 (1.2) |
|  | Missing | 22 (1.4) | 53 (1.4) | 11 (1.7) | 17 (1.1) |
| rs10755709 | A/A | 708 (45.9) | 1,824 (48.1) | 300 (45.4) | 749 (46.1) |
|  | G/A | 618 (40.0) | 1,535 (40.5) | 291 (44.0) | 701 (43.1) |
|  | G/G | 169 (11.0) | 332 (8.8) | 58 (8.8) | 124 (7.6) |
|  | Missing | 49 (3.2) | 100 (2.6) | 12 (1.8) | 51 (3.1) |
| rs16873740 | T/T | 1,171 (75.8) | 2,972 (78.4) | 495 (74.9) | 1,266 (77.9) |
|  | A/T | 340 (22.0) | 763 (20.1) | 157 (23.8) | 336 (20.7) |
|  | A/A | 32 (2.1) | 52 (1.4) | 8 (1.2) | 20 (1.2) |
|  | Missing | 1 (0.1) | 4 (0.1) | 1 (0.2) | 3 (0.2) |
|  |  | Cases<br>N=1,544 | Controls<br>N=3,791 | Cases<br>N=661 | Controls<br>N=1,625 |
| rs118072448 | T/T | 1,346 (87.2) | 3,215 (84.8) | 581 (87.9) | 1,380 (84.9) |
|  | C/T | 188 (12.2) | 536 (14.1) | 71 (10.7) | 231 (14.2) |
|  | C/C | 10 (0.7) | 40 (1.1) | 9 (1.4) | 14 (0.9) |
|  | Missing | 0 (0.0) | 0 (0.0) | 0 (0.0) | 0 (0.0) |
| rs7027911 | G/G | 367 (23.8) | 1,023 (27.0) | 156 (23.6) | 398 (24.5) |
|  | A/G | 606 (39.3) | 1,415 (37.3) | 261 (39.5) | 676 (41.6) |
|  | A/A | 240 (15.5) | 553 (14.6) | 111 (16.8) | 229 (14.1) |
|  | Missing | 331 (21.4) | 800 (21.1) | 133 (20.1) | 322 (19.8) |
| rs71481792 | A/A | 239 (15.5) | 514 (13.6) | 122 (18.5) | 246 (15.1) |
|  | T/A | 701 (45.4) | 1,704 (45.0) | 263 (39.8) | 712 (43.8) |
|  | T/T | 522 (33.8) | 1,416 (37.4) | 247 (37.4) | 591 (36.4) |
|  | Missing | 82 (5.3) | 157 (4.1) | 29 (4.4) | 76 (4.7) |
| rs1984162 | A/A | 827 (53.6) | 2,144 (56.6) | 363 (54.9) | 865 (53.2) |
|  | G/A | 612 (39.6) | 1,416 (37.4) | 249 (37.7) | 642 (39.5) |
|  | G/G | 105 (6.8) | 231 (6.1) | 49 (7.4) | 118 (7.3) |
|  | Missing | 0 (0.0) | 0 (0.0) | 0 (0.0) | 0 (0.0) |
| rs115492982 | G/G | 1,529 (99.0) | 3,774 (99.6) | 654 (98.9) | 1,621 (99.8) |
|  | A/G | 14 (0.9) | 15 (0.4) | 7 (1.1) | 3 (0.2) |
|  | A/A | 1 (0.1) | 0 (0.0) | 0 (0.0) | 0 (0.0) |
|  | Missing | 0 (0.0) | 2 (0.1) | 0 (0.0) | 1 (0.1) |
|  |  | Cases<br>N=1,544 | Controls<br>N=3,791 | Cases<br>N=661 | Controls<br>N=1,625 |
| rs112317747 | T/T | 1,410 (91.3) | 3,518 (92.8) | 610 (92.3) | 1,521 (93.6) |
|  | C/T | 115 (7.5) | 236 (6.2) | 44 (6.7) | 87 (5.4) |
|  | C/C | 2 (0.1) | 0 (0.0) | 0 (0.0) | 0 (0.0) |
|  | Missing | 17 (1.1) | 37 (1.0) | 7 (1.1) | 17 (1.1) |
| rs2034831 | A/A | 1,284 (83.2) | 3,242 (85.5) | 550 (83.2) | 1,375 (84.6) |
|  | C/A | 200 (13.0) | 399 (10.5) | 80 (12.1) | 190 (11.7) |
|  | C/C | 8 (0.5) | 18 (0.5) | 10 (1.5) | 8 (0.5) |
|  | Missing | 52 (3.4) | 132 (3.5) | 21 (3.2) | 52 (3.2) |
| rs35896106 | C/C | 1,251 (81.0) | 3,166 (83.5) | 537 (81.2) | 1,373 (84.5) |
|  | T/C | 231 (15.0) | 514 (13.6) | 104 (15.7) | 203 (12.5) |
|  | C/C | 13 (0.8) | 23 (0.6) | 5 (0.8) | 12 (0.7) |
|  | Missing | 49 (3.2) | 88 (2.3) | 15 (2.3) | 37 (2.3) |
| rs76374459 | G/G | 1,318 (85.4) | 3,320 (87.6) | 567 (85.8) | 1,440 (88.6) |
|  | C/G | 187 (12.1) | 394 (10.4) | 83 (12.6) | 150 (9.2) |
|  | C/C | 9 (0.6) | 12 (0.3) | 1 (0.2) | 8 (0.5) |
|  | Missing | 30 (1.9) | 65 (1.7) | 10 (1.5) | 27 (1.7) |
| rs35652899 | C/C | 1,286 (83.3) | 3,236 (85.4) | 553 (83.7) | 1,406 (86.5) |
|  | G/C | 222 (14.4) | 493 (13.0) | 97 (14.7) | 187 (11.5) |
|  | G/G | 14 (0.9) | 20 (0.5) | 3 (0.5) | 10 (0.6) |
|  | Missing | 22 (1.4) | 42 (1.1) | 8 (1.2) | 22 (1.4) |
|  |  | Cases<br>N=1,544 | Controls<br>N=3,791 | Cases<br>N=661 | Controls<br>N=1,625 |
| rs76488148 | G/G | 1,385 (89.7) | 3,463 (91.4) | 603 (91.2) | 1,488 (91.6) |
|  | T/G | 144 (9.3) | 290 (7.7) | 49 (7.4) | 119 (7.3) |
|  | T/T | 5 (0.3) | 7 (0.2) | 0 (0.0) | 4 (0.3) |
|  | Missing | 10 (0.7) | 31 (0.8) | 9 (1.4) | 14 (0.9) |

SD, standard deviation

### Validation of prototype model

For the validation of our prototype risk model [9], we used the 1234 cases and 4805 controls that were not included in our previous paper. We constructed relative risk scores for both the clinical model and the combined clinical and SNP score model using the exponent of the sum of the intercept and the beta coefficients for each risk factor in the prototype model [9].

### Development and validation of the new model

To develop a new model to predict risk of severe COVID-19, we used all of the available data and randomly divided it into a 70% training dataset and a 30% validation dataset (ensuring that the datasets were balanced for case and control status). We used multiple imputation with 20 imputations to address the missing data for body mass index (linear regression) and the SNP data (predictive mean matching) for the development of the new model in the training dataset. To more closely reflect the availability of data in the real world, we did not use imputed data in the validation dataset.

The clinical variables considered for inclusion in the new model were age, sex, body mass index (BMI), ethnicity (Caucasian vs other), ABO blood type and the following chronic health conditions: asthma, autoimmune disease (rheumatoid arthritis, lupus or psoriasis), haematological cancer, non-haematological cancer, cerebrovascular disease, diabetes, heart disease, hypertension, immunocompromised, kidney disease, liver disease and respiratory disease (excluding asthma). Dummy variables were used for the categorical classifications of age and ABO blood type.

The SNPs selected for consideration in the development of the new model came from three sources: (i) the 64 SNPs from our prototype model [9], (ii) the 12 SNPs from the paper by Pairo-Castineira et al. [14], and (iii) the 40 SNPs newly selected from the results of the COVID-19 Host Genetics Initiative’s COVID19-hg GWAS meta-analyses round 4 meta-analysis of non-hospitalized versus hospitalized cases of COVID-19 [10, 22]. To avoid reliance on potentially inaccurate summary statistics to construct a polygenic risk score, we used unadjusted logistic regression in the multiple imputation training dataset to identify the subset of SNPS that were associated with risk of severe COVID-19 with P<0.05 (see Supplementary Table S1) and used these as individual risk factors (with a per allele effect) to build our new model.

### Statistical methods

#### Development of new model

We used multivariable logistic regression in the multiple imputation training dataset to develop the new model to predict risk of severe COVID-19. We began with a model that included all of the clinical variables and the SNPs with unadjusted associations with severe COVID-19. We then used backwards stepwise selection to develop the most parsimonious model. For the removed variables we made a final determination on their inclusion or exclusion by adding them one at a time to the parsimonious model. To directly compare the effect sizes of the variables in the final model, regardless of the scale on which they were measured, we used the odds per adjusted standard deviation [23]. We used the intercept and beta coefficients from the new model to calculate the COVID-19 risk score (as a % risk) for all eligible UK Biobank participants.

#### Model performance

The association between risk score and severe COVID-19 was assessed using logistic regression to estimate the OR per quintile of risk score. We assessed model discrimination using the area under the receiver operating characteristic curve (AUC). Where warranted, we plotted the receiver operating characteristic curve of the model.

We assessed calibration using logistic regression of the log odds of the risk score to estimate the intercept and the slope (beta coefficient). An intercept close to 0 indicates good calibration, while an intercept of less than 0 indicates overall overestimation and an intercept of greater than 0 indicates overall underestimation of risk.

In terms of the dispersion of the risk score, a slope of close to 1 indicates good estimation across the spectrum of risk. A slope of less than 1 means that the predicted probabilities do not vary enough (i.e. underestimation of true high risk and overestimation of true low risk). Conversely, a slope of greater than one means that the predicted probabilities vary too much (i.e. underestimation of true low risk and overestimation of true high risk). Where helpful, we also used a calibration plot to illustrate the fit of a model.

We used Stata (version 16.1) [24] for analyses; all statistical tests were two-sided and P<0.05 was considered nominally statistically significant.

### Ethics approval

The UK Biobank has Research Tissue Bank approval (REC #11/NW/0382) that covers analysis of data by approved researchers. All participants provided written informed consent to the UK Biobank before data collection began. This research has been conducted using the UK Biobank resource under Application Number 47401.

### Data availability statement

The data underlying this article was provided by the UK Biobank and we do not have permission to share the data. Researchers wishing to access the data used in this study can apply directly to the UK Biobank at https://www.ukbiobank.ac.uk/register-apply/. Stata 16.1 code for the analysis is available from the corresponding author on request.

## Results

In the results file downloaded on 8 January 2021, there were 2205 eligible cases with severe COVID-19 and 5416 eligible controls with non-severe COVID-19.

### Validation of prototype model

Characteristics of the new UKB participants (1234 cases and 4805 controls) with positive SARS-CoV-2 test results are shown in Supplementary Table S2.

The odds ratio (OR) per quintile showed that the clinical risk score was strongly associated with severe COVID-19 (OR=1.70; 95% confidence interval [CI]=1.62, 1.79; P<0.001) and that the combined clinical and SNP risk score was less strongly associated with severe COVID-19 (OR=1.45; 95% CI=1.38, 1.52; P<0.001); there was no association with severe COVID-19 for the SNP score (OR=0.98; 95% CI=0.94, 1.03; P=0.5). The discrimination of cases and controls was excellent for the clinical score (AUC=0.711; 95% CI=0.694, 0.727), lower for the combined clinical and SNP score (AUC=0.657; 95% CI=0.639, 0.674) and poor for the SNP score alone (AUC=0.491; 95% CI=0.473, 0.509).

Assessment of model calibration showed that overall, risk was overestimated for both the clinical risk model (α=−1.72; 95% CI=−1.80, −1.65; P<0.001) and the clinical and SNP model (α=−1.63; 95% CI=−1.71, −1.54; P<0.001). For the clinical model, there was no evidence of poor dispersion (β=1.03, 95% CI=0.94, 1.12, P=0.5), while the predictions of the combined clinical and SNP model varied too much (β=0.59, 95% CI=0.52, 0.65, P<0.001).

### Development and validation of the new model

Table 1 shows the characteristics of the 1544 cases and 3791 controls in the 70% training dataset and the 661 cases and 1,625 controls in the 30% validation data set. In the training dataset, the mean age was 69.8 years (SD=8.6) for cases and 64.6 years (SD=8.4) for controls, and the mean BMI was 29.3 kg/m^2^ (SD=5.3) for cases and 28.0 kg/m^2^ (SD=4.9) for controls. In the validation dataset, the mean age was 69.7 years (SD=8.7) for cases and 64.4 years (SD=8.4) for controls, and the mean BMI was 29.4 kg/m^2^ (SD=5.6) for cases and 28.3 kg/m^2^ (SD=5.0) for controls.

#### Training

In the age and sex model, being male and being in one of the four older age groups conferred a substantially increased risk of severe COVID-19 (Table 2), with an OR=1.60 for being male and ORs ranging from 2.74 for the age groups from 65–69 years to 4.95 for the 80+ years group. Direct comparison of the effect size of each variable showed that the age group 75–79 years was the strongest risk factor (with odds per adjusted standard deviation of 1.58), followed by the 70–74 and 80–84 groups (with odds per adjusted standard deviations of 1.42 and 1.34, respectively).

**Table 2.** Age and sex model for risk of severe COVID-19 in the training dataset.

| Variable |  | Adjusted odds ratio | 95% confidence interval | P value | Odds per adjusted standard deviation | 95% confidence interval |
| --- | --- | --- | --- | --- | --- | --- |
| Age group (years) | 65–69 | 1.60 | 1.32, 1.94 | <0.001 | 1.18 | 1.10, 1.26 |
|  | 70–74 | 2.74 | 2.31, 3.24 | <0.001 | 1.42 | 1.34, 1.50 |
|  | 75–79 | 4.20 | 3.55, 4.97 | <0.001 | 1.58 | 1.50, 1.67 |
|  | 80+ | 4.95 | 3.83, 6.39 | <0.001 | 1.34 | 1.28, 1.41 |
| Sex | Male | 1.48 | 1.31, 1.67 | <0.001 | 1.21 | 1.14, 1.29 |

Note: Adjusted odds ratio and odds per adjusted standard deviation calculated using the original dataset because no missing data in this model.

The new model was developed from the variables in Table 1, which include the clinical variables and the 14 SNPs identified as having unadjusted per allele ORs with P-values <0.05 (see Supplementary Table S1). The variables retained in the new model are shown in Table 3 and comprise three age groups (70–74, 75–79 and 80–84 years), sex, ethnicity, body mass index, six comorbidities and seven SNPs. Compared with the age and sex model, the effects of sex and age group were attenuated in the new model, with an OR=1.27 for being male, the 70–74 years age group not being at increased risk, and ORs for the other age groups ranging from 1.77 for the 70–74 years group to 2.76 for the 80+ years group.

**Table 3.** Adjusted odds ratios and odds per adjusted standard deviations for the risk factors in the new model for risk of severe COVID-19 in the training dataset.

| Variable |  | Adjusted odds ratio | 95% confidence interval | P value | Odds per adjusted standard deviation | 95% confidence interval |
| --- | --- | --- | --- | --- | --- | --- |
| Age group (years) | 70–74 | 1.77 | 1.49, 2.12 | <0.001 | 1.22 | 1.15, 1.30 |
|  | 75–79 | 2.28 | 1.90, 2.73 | <0.001 | 1.29 | 1.22, 1.36 |
|  | 80+ | 2.76 | 2.09, 3.64 | <0.001 | 1.20 | 1.14, 1.26 |
| Sex | Male | 1.27 | 1.12, 1.46 | <0.001 | 1.13 | 1.06, 1.20 |
| Ethnicity | Non-white | 1.34 | 1.06, 1.70 | 0.02 | 1.08 | 1.01, 1.14 |
| Inverse of body mass index | 10/(kg/m<sup>2</sup>) | 0.20 | 0.06, 0.66 | 0.008 | 0.91 | 0.85, 0.97 |
| Cancer – haematological | Yes | 2.73 | 1.62, 4.60 | <0.001 | 1.09 | 1.04, 1.13 |
| Cancer – non-haematological | Yes | 1.29 | 1.08, 1.54 | 0.005 | 1.09 | 1.03, 1.15 |
| Cerebrovascular disease | Yes | 1.50 | 1.17, 1.92 | 0.001 | 1.08 | 1.03, 1.14 |
| Diabetes | Yes | 1.54 | 1.26, 1.87 | <0.001 | 1.12 | 1.06, 1.18 |
| Hypertension | Yes | 1.34 | 1.15, 1.56 | <0.001 | 1.12 | 1.06, 1.19 |
| Kidney disease | Yes | 2.00 | 1.53, 2.61 | <0.001 | 1.12 | 1.07, 1.17 |
| Respiratory disease (excluding asthma) | Yes | 3.23 | 2.71, 3.85 | <0.001 | 1.35 | 1.29, 1.42 |
| rs112641600 | Per T allele | 0.79 | 0.68, 0.92 | 0.003 | 0.90 | 0.84, 0.97 |
| rs10755709 | Per G allele | 1.13 | 1.02, 1.25 | 0.02 | 1.09 | 1.02, 1.16 |
| rs118072448 | Per C allele | 0.82 | 0.69, 0.98 | 0.03 | 0.93 | 0.86, 0.99 |
| rs7027911 | Per A allele | 1.11 | 1.00, 1.23 | 0.05 | 1.07 | 1.00, 1.15 |
| rs71481792 | Per T allele | 0.90 | 0.82, 1.00 | 0.04 | 0.93 | 0.87, 0.99 |
| rs112317747 | Per C allele | 1.31 | 1.02, 1.70 | 0.04 | 1.06 | 1.00, 1.13 |
| rs2034831 | Per C allele | 1.27 | 1.05, 1.53 | 0.01 | 1.08 | 1.02, 1.15 |

Note: Using multiple imputation data; odds per adjusted standard deviation calculated using only the first imputation dataset.

Direct comparison of the effect size of each variable showed that respiratory disease was the strongest risk factor with odds per adjusted standard deviation of 1.35, followed by the three older age groups with odds per adjusted standard deviations of 1.20 to 1.29). The other clinical risk factors and SNPs had odds per adjusted standard deviation in the range 1.07 to 1.13 (or the equivalent protective effect).

The age and sex model had good discrimination of cases and controls with an AUC of 0.676 (95% CI=0.659, 0.692) but the new model with an AUC of 0.752 (95% CI=0.737, 0.767) was a substantial improvement (χ^2^=149.40, df=1, P<0.001).

#### Validation

In the non-imputed validation dataset, the age and sex model and the new model were associated with severe COVID-19. The OR per quintile for the age and sex model was 1.49 (95% CI=1.40, 1.59; P<0.001), while the new model had a substantially higher OR per quintile of 1.77 (95% CI=1.64, 1.90; P<0.001).

In terms of discrimination between cases and controls, the age and sex model had an AUC of 0.671 (95% CI=0.646, 0.696), while the new model with an AUC of 0.732 (95% CI=0.708, 0.756) was a substantial improvement (χ^2^=41.23, df=1, P<0.001). The receiver operating characteristic curves for both models are shown in Figure 1.

**Figure 1.**
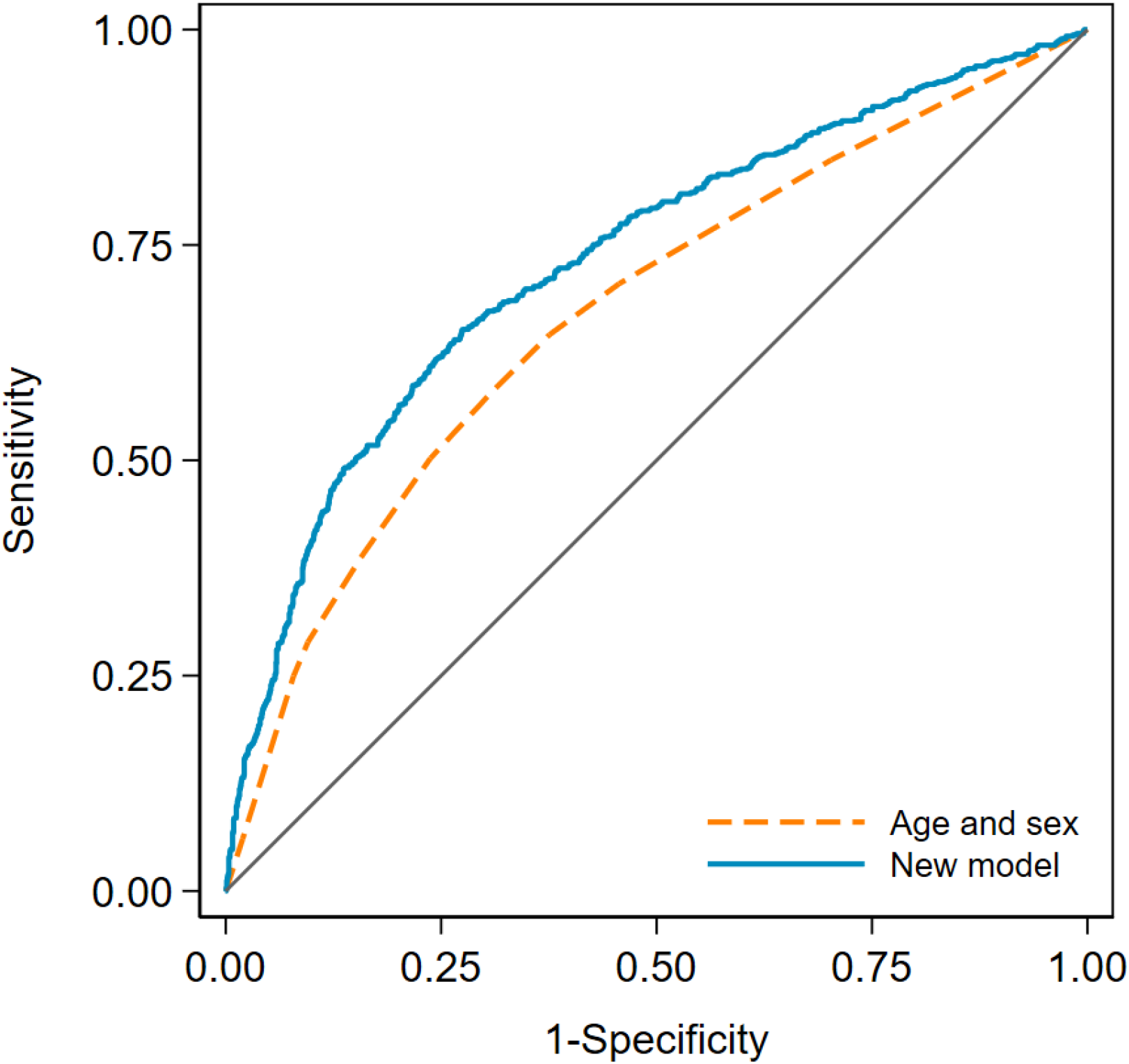
Receiver operating characteristic curves for the age and sex model and the new model in the validation dataset. The new model has an area under the curve (AUC) of 0.732 (95% CI=0.708, 0.756), and the age and sex model has an AUC of 0.671 (95% CI=0.646, 0.696).

Both models were well calibrated with no evidence of overall overestimation or underestimation for the age and sex model (α=−0.02; 95% CI=−0.18, 0.13; P=0.7) or the new model (α=−0.08; 95% CI=−0.21, 0.05; P=0.3). There was also no evidence of under or over dispersion for the age and sex model (β=0.96, 95% CI=0.81, 1.10, P=0.6) and for the new model (β=0.90, 95% CI=0.80, 1.00, P=0.06). Calibration plots for both models are shown in Figure 2.

**Figure 2.**
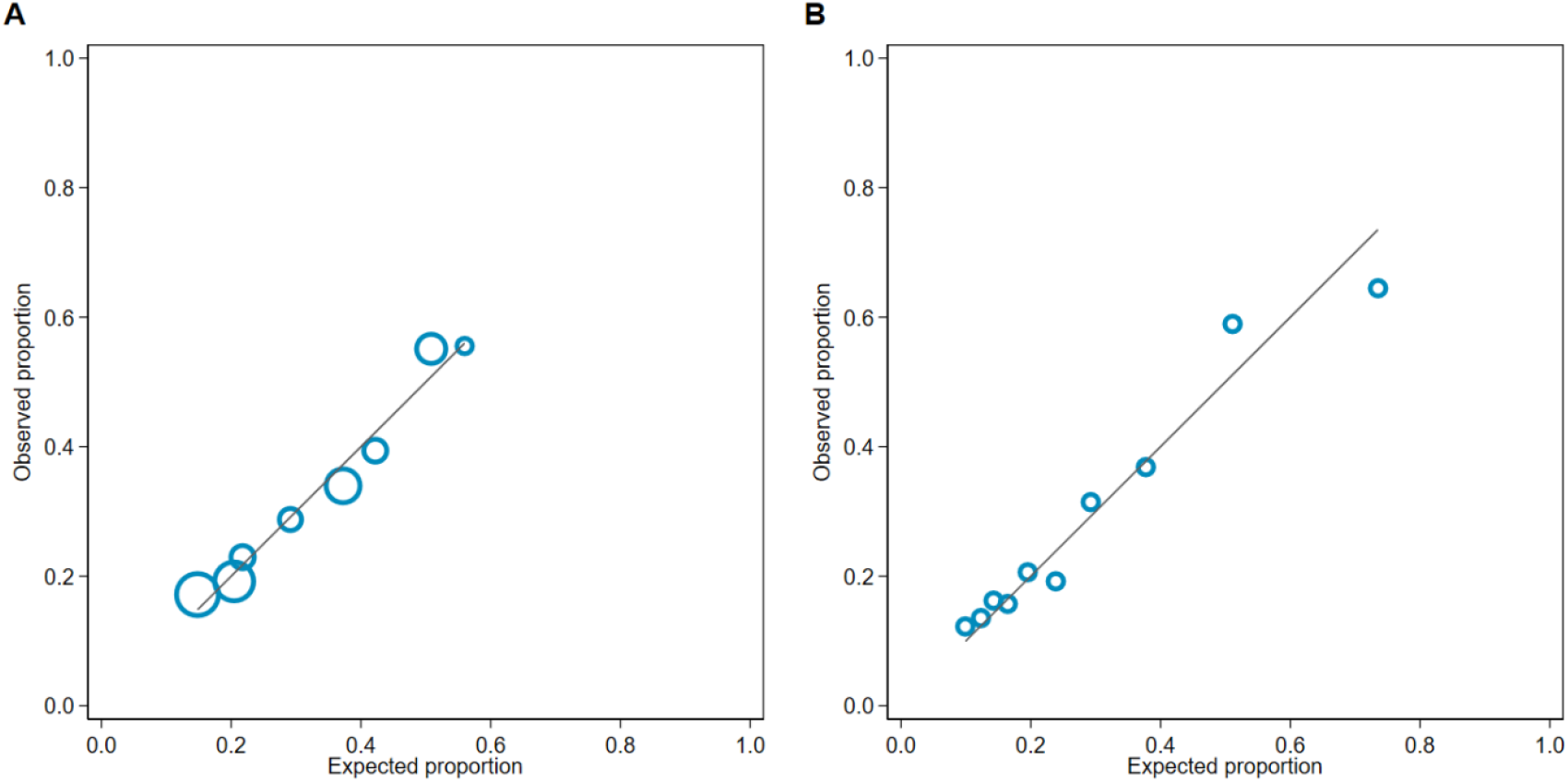
Calibration plots for the (A) age and sex model and (B) new model in the validation dataset.

#### Probability of severe COVID-19 in whole UK Biobank

We calculated the probability of severe COVID-19 for all UK Biobank participants who met our eligibility criteria for this study; the distributions are shown in Figure 3, and the distribution of the new model by 5-year age group are shown in Supplementary Figure S1. Using the age and sex model, the mean probability was 0.32 (SD=0.13) and ranged from a minimum of 0.15 to a maximum of 0.56. Using the new model, the mean probability was 0.27 (SD=0.16) and the range was from 0.04 to 0.98, a much wider range than for the age and sex model.

**Figure 3.**
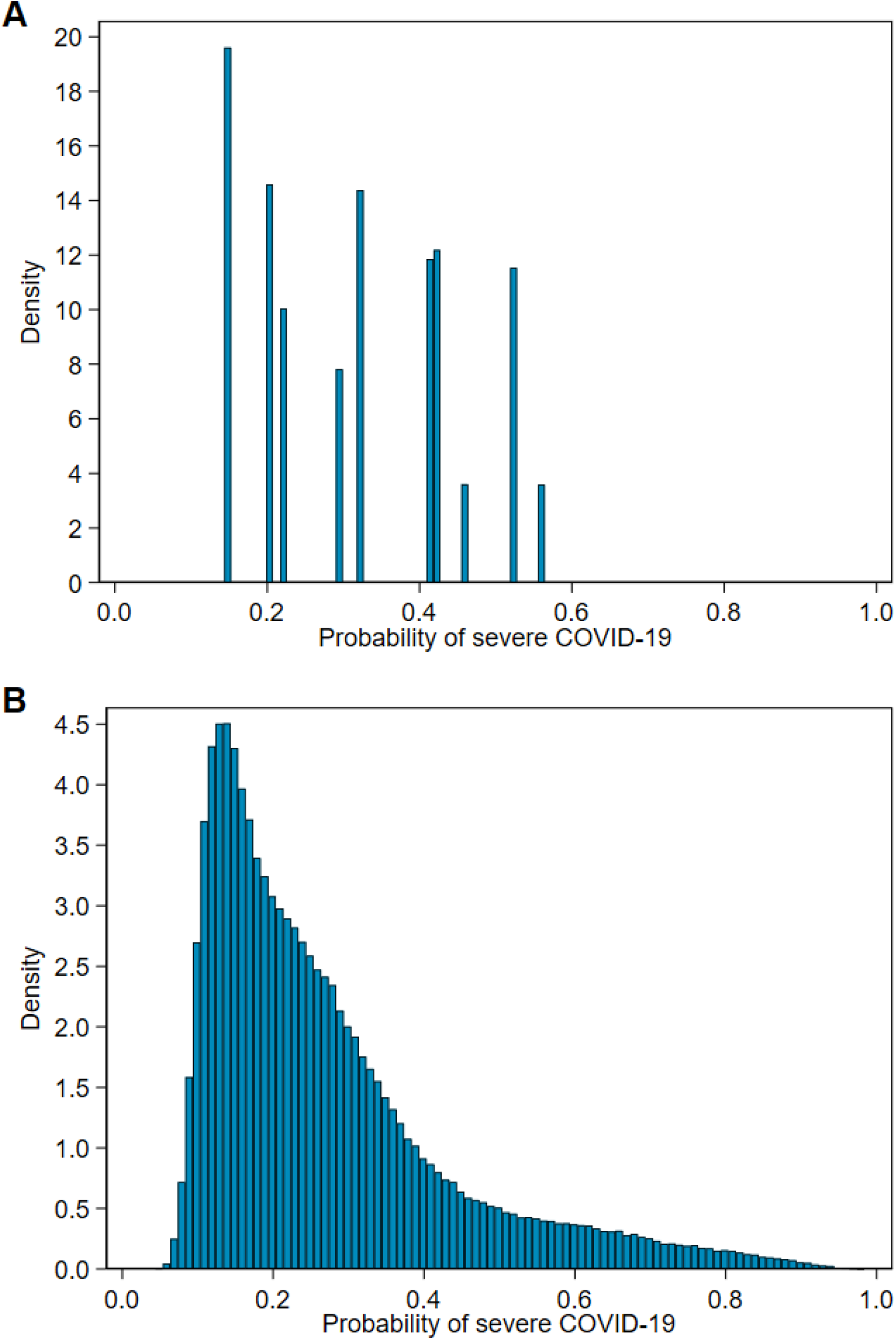
Distribution of probability of severe COVID-19 in all of UK Biobank for (A) the age and sex model and (B) the new model.

## Discussion

An accurate test to predict risk of severe COVID-19 can inform prioritization of vaccine doses to those most at risk [25] and will be useful in regions in which vaccination is not widespread enough to provide herd immunity (either through unavailability or vaccine hesitancy), if available vaccines are not effective against variants of SARS-CoV-2, or if available vaccines are not indicated for some people. On an individual level, knowledge of personal risk can empower people to make informed choices about their day-to-day activities, including targeted social distancing in the workplace [26] or other crowded places.

The validation of the clinical component of our prototype model confirmed that it performed well with good discrimination (AUC=0.711), but overall, it overestimated risk. The SNP score component of the prototype model was not confirmed in the validation dataset and is likely due to the prototype model having been developed in dataset with a high prevalence of severe COVID-19.

Given the failure to confirm our prototype SNP-based risk score, we incorporated SNPs in the new model without relying on published summary statistics and without assumptions as to the identity of the risk allele. We included the SNPs as individual risk factors and estimated the per allele OR for each. By doing so, we were able to identify the subset of SNPs and clinical risk factors that were informative for predicting risk. These risk factors are all important to risk prediction, and characterization of the SNP genotypes is as important as ascertaining clinical information.

From our initial list of 116 SNPs (Supplementary Table S1), we considered 14 for inclusion in our model and retained seven, none of which were in the 3p21.31 locus identified by others [12-14, 22]. Functionally, most of the SNPs retained in our new model are associated with genes that play a role in infection pathways or immunity. The immune function and chromatin remodelling family of GATA transcription factors are associated by the inclusion of SNPs near *HIVEP1* (rs10755709), which encodes a viral-infection regulation transcription factor, and *GATA3* (rs71481792) [27, 28]. *ALPK1* and *TIFA* are closely downstream of rs112641600 and both have adaptive and innate signal transduction roles and pro-inflammatory functions [29]. *MSR1*, upstream of rs118072448, is a macrophage scavenger receptor and implicated in a broad range of disease types including host viral defence [30] and *PSAT1* is associated with glutamine metabolic reprogramming by SARS-CoV-2 and viral mRNA translation [31].

In the development of the new model, the strongest risk factor was respiratory disease (with an odds per standard deviation of 1.35; Table 3). The older age groups (70–74, 75–79, and 80+ years) and being male all had odds per standard deviations of 1.20 to 1.29. The other risk factors (the seven SPNs, ethnicity, body mass index, cancer history (haematological and non-haematological), cerebrovascular disease, diabetes, hypertension, and kidney disease) all had odds per adjusted standard deviations in the range 1.07 to 1.13 (or the equivalent protective effect).

In the non-imputed validation dataset, the new model performed very well with an AUC of 0.732 (compared with an AUC of 0.752 in the training dataset). Importantly, the new model was well calibrated, showing no evidence of problems with the overall estimation of risk or the dispersion of risk predictions. The validation of the new model also illustrates the importance of considering risk factors beyond age and sex in predicting risk of severe COVID-19. The new model was a substantial improvement over the age and sex model, in terms of the OR per quintile (OR=1.77 and OR=1.49, respectively) and the discrimination of cases and controls (AUC=0.732 and AUC=0.671, respectively). The new model also allows stratification across a wide range of risk (Figure 3B) so that, for example, a healthy person aged 75 years might have a lower risk of severe COVID-19 than a 50-year-old person with several risk factors.

A limitation of this study is that, through necessity, we used hospitalization as a proxy for COVID-19 severity and the outcome measure may have been misclassified for some participants. This would have attenuated the observed associations and it is possible that some risk factors have been omitted unnecessarily. Nevertheless, we are confident in the variables retained. We were also unable to develop models for other important endpoints such as intensive care admission or death.

The progression of the COVID-19 pandemic has seen people experience chronic symptoms, and some of these people will have had only a mild original infection [5]. Identifying people who are at increased risk of chronic disease is an obvious direction for future research. Another direction for future research is to investigate whether our model for the prediction of severe COVID-19 is applicable for the new SARS-CoV-2 variants of concern, which have been reported to have increased transmissibility, virulence and antigenicity and cause more severe disease [3, 4]. Further validation of our new model is required in independent datasets, especially those in which the SARS-CoV-2 variant has been characterized.

Clear benefits of our new model for predicting risk of severe COVID-19 are that the required clinical data is simple to collect and that the genetic information is amenable to high-throughput genotyping, with rapid turnaround that is essential for the present pandemic. In the light of the uncertainty of the future of the COVID-19 pandemic, accurate knowledge of individual risk of severe COVID-19 can make an important contribution to healthcare on both a population and a personal level.

## Data Availability

We used data from the UK Biobank (under Application Number 47401) for these analyses and do not have permission to share the data. Researchers wishing to access the data used in this study can apply directly to the UK Biobank at https://www.ukbiobank.ac.uk/register-apply/.

## Acknowledgements

We wish to thank Mr Lawrence Whiting for his invaluable expertise in the management of large data files from the UK Biobank.

## Supplementary Information

### Supplementary figure

**Supplementary Figure S1.**
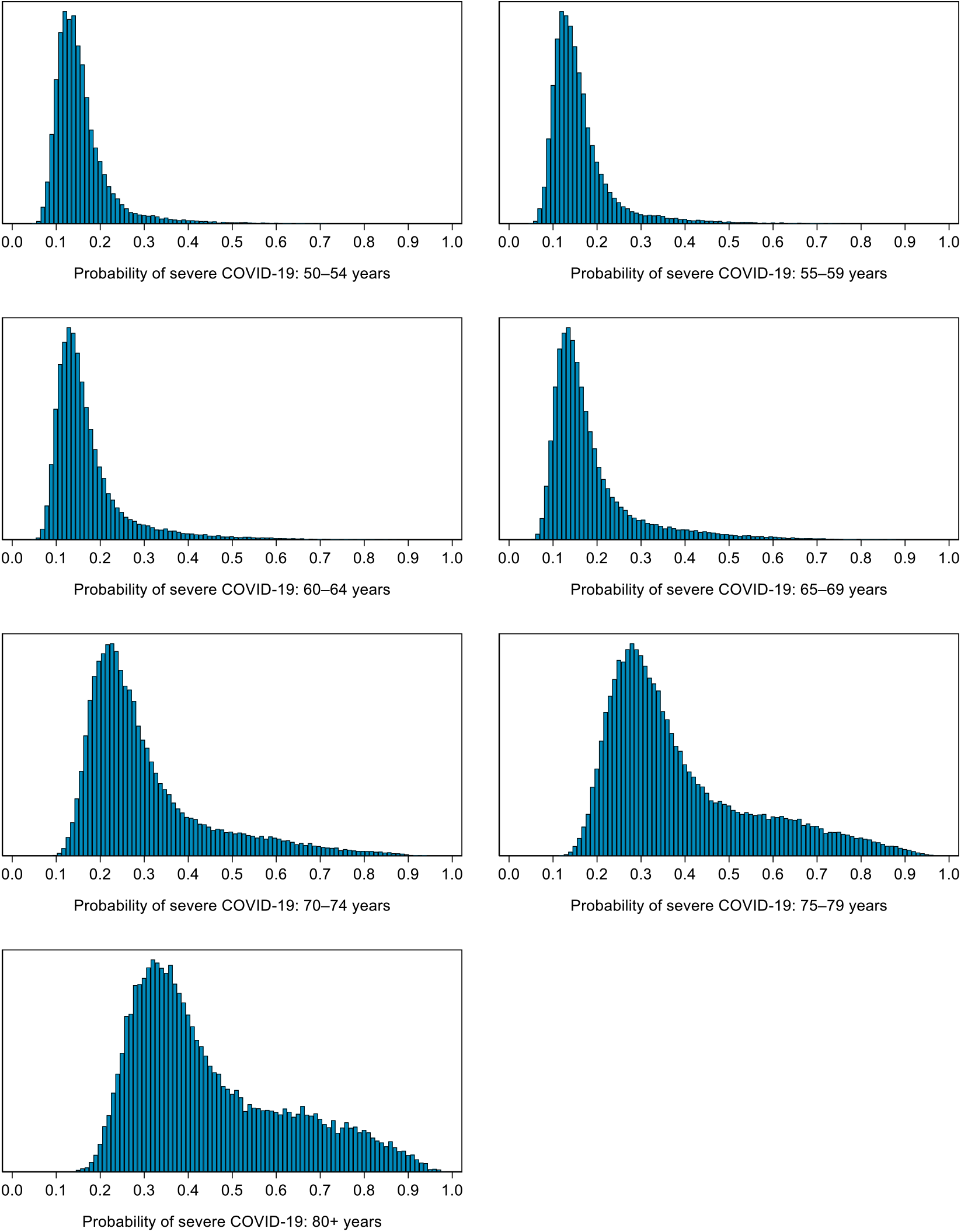
Distribution of probability of severe COVID-19 by 5-year age group.

### Supplementary tables

**Supplementary Table S1.**
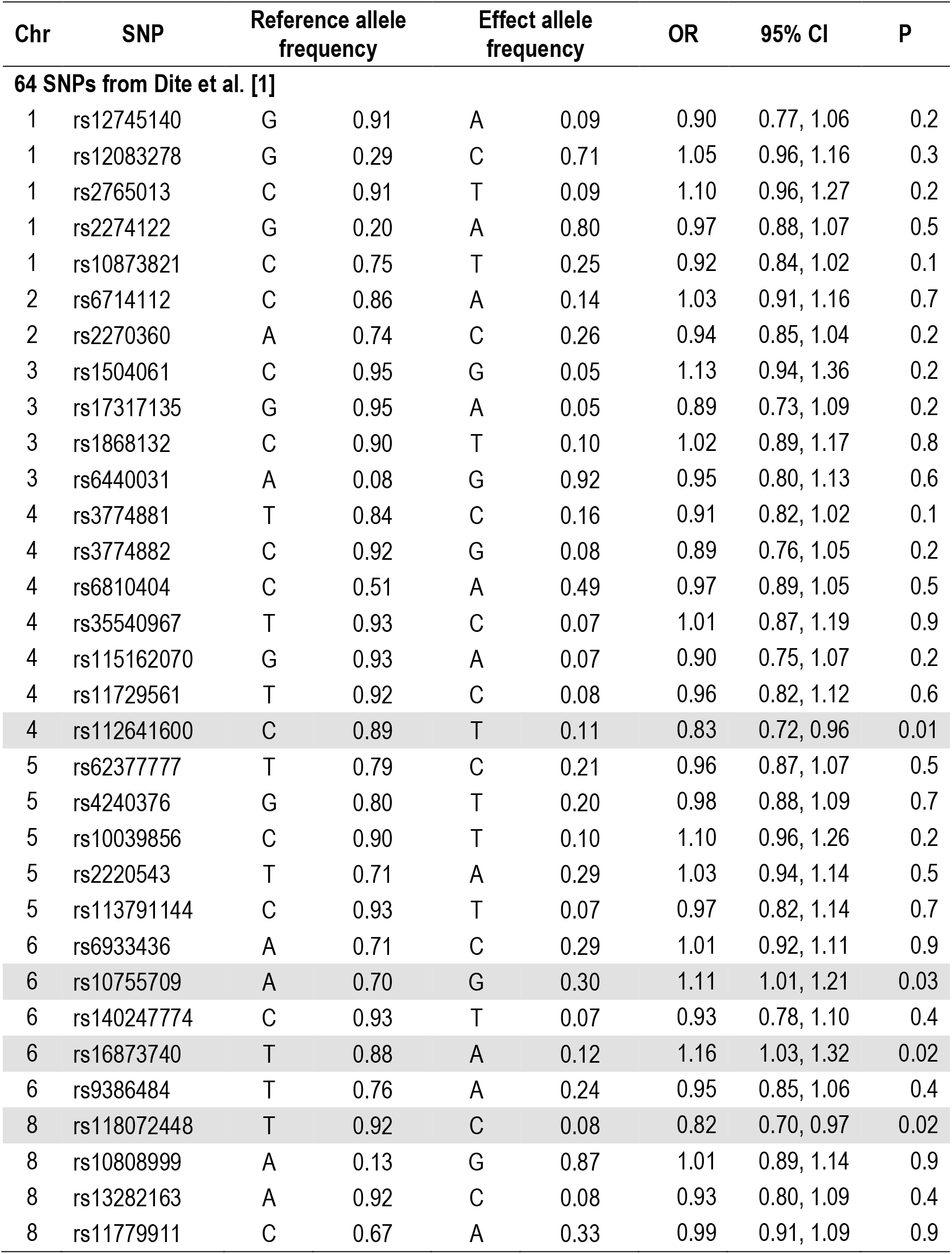
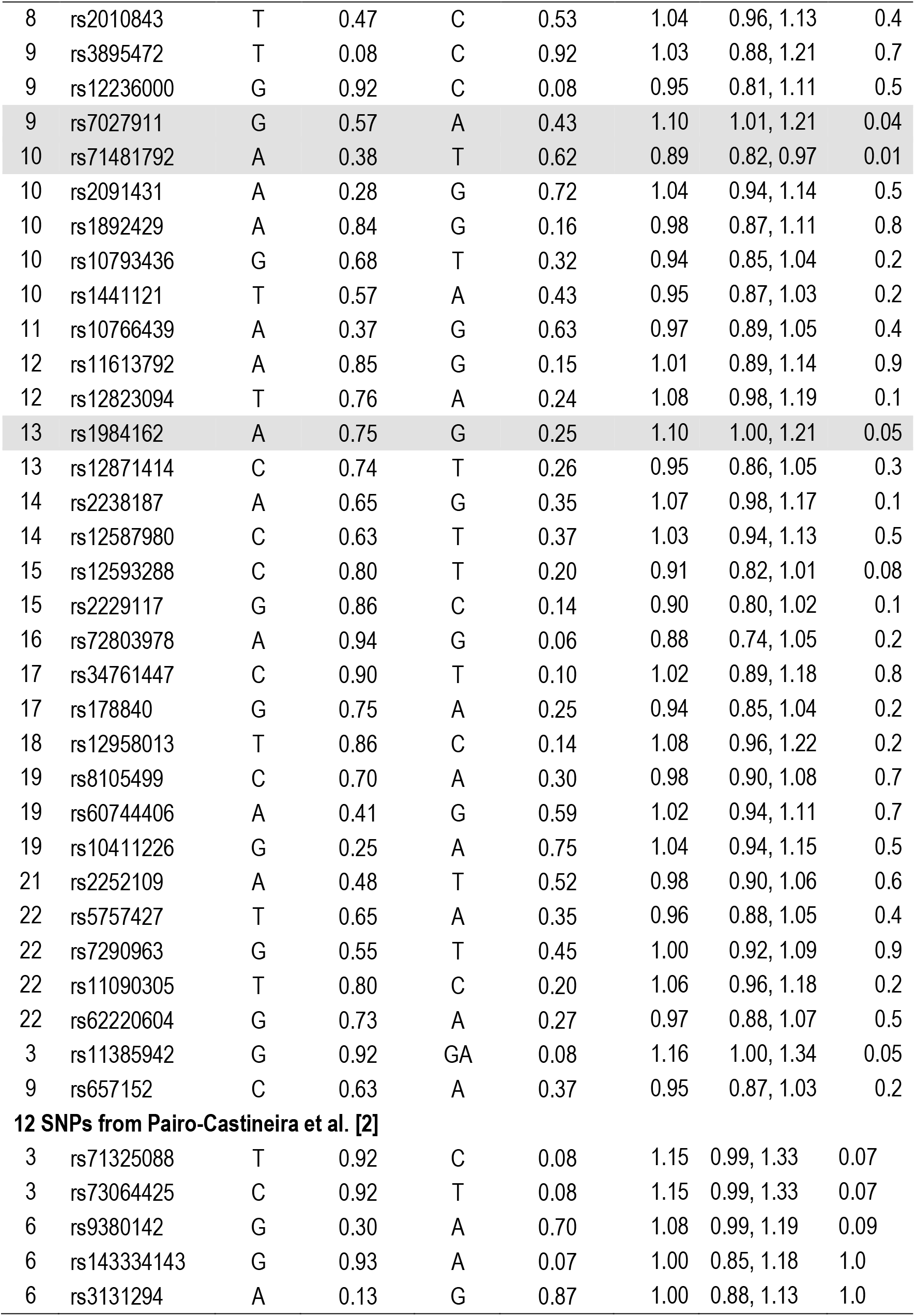
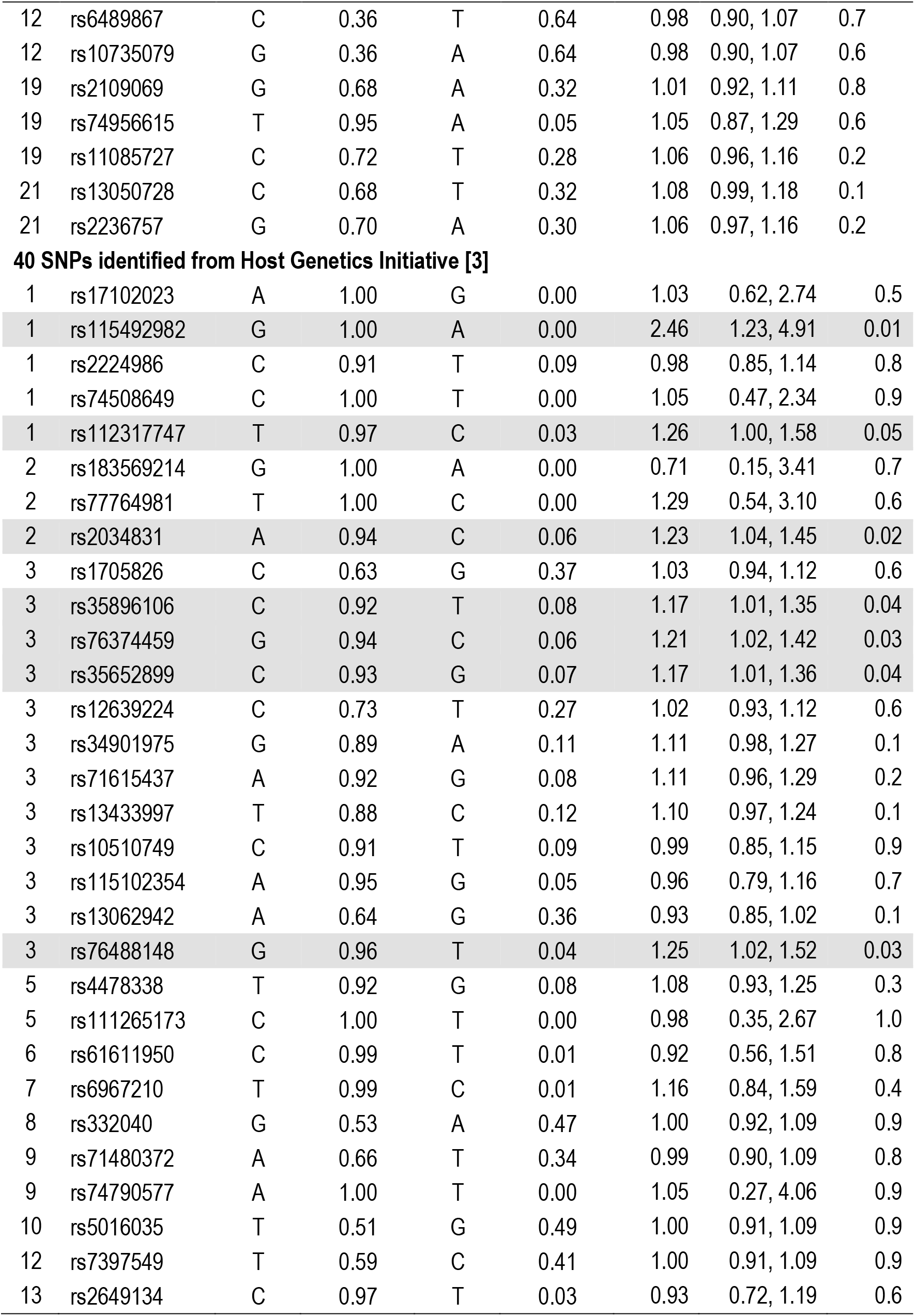
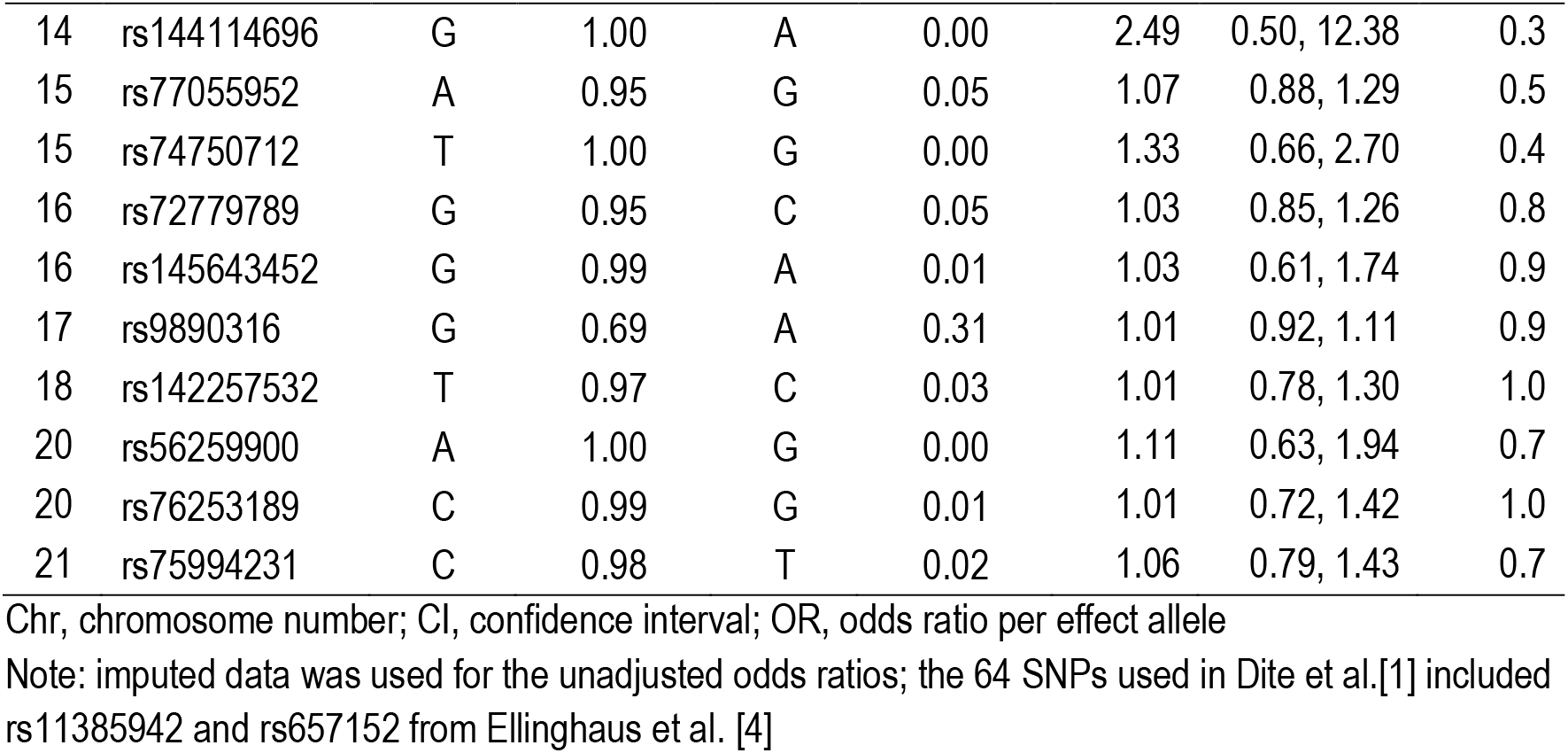
Allele frequencies and unadjusted odds ratios for the full list of SNPs identified as potential risk factors for severe COVID-19 – training dataset (shaded SNPs were selected)

| Chr | SNP | Reference allele frequency |  | Effect allele frequency |  | OR | 95% CI | P |
| --- | --- | --- | --- | --- | --- | --- | --- | --- |
| 64 SNPs from Dite et al. [1] |  |  |  |  |  |  |  |  |
| 1 | rs12745140 | G | 0.91 | A | 0.09 | 0.90 | 0.77, 1.06 | 0.2 |
| 1 | rs12083278 | G | 0.29 | C | 0.71 | 1.05 | 0.96, 1.16 | 0.3 |
| 1 | rs2765013 | C | 0.91 | T | 0.09 | 1.10 | 0.96, 1.27 | 0.2 |
| 1 | rs2274122 | G | 0.20 | A | 0.80 | 0.97 | 0.88, 1.07 | 0.5 |
| 1 | rs10873821 | C | 0.75 | T | 0.25 | 0.92 | 0.84, 1.02 | 0.1 |
| 2 | rs6714112 | C | 0.86 | A | 0.14 | 1.03 | 0.91, 1.16 | 0.7 |
| 2 | rs2270360 | A | 0.74 | C | 0.26 | 0.94 | 0.85, 1.04 | 0.2 |
| 3 | rs1504061 | C | 0.95 | G | 0.05 | 1.13 | 0.94, 1.36 | 0.2 |
| 3 | rs17317135 | G | 0.95 | A | 0.05 | 0.89 | 0.73, 1.09 | 0.2 |
| 3 | rs1868132 | C | 0.90 | T | 0.10 | 1.02 | 0.89, 1.17 | 0.8 |
| 3 | rs6440031 | A | 0.08 | G | 0.92 | 0.95 | 0.80, 1.13 | 0.6 |
| 4 | rs3774881 | T | 0.84 | C | 0.16 | 0.91 | 0.82, 1.02 | 0.1 |
| 4 | rs3774882 | C | 0.92 | G | 0.08 | 0.89 | 0.76, 1.05 | 0.2 |
| 4 | rs6810404 | C | 0.51 | A | 0.49 | 0.97 | 0.89, 1.05 | 0.5 |
| 4 | rs35540967 | T | 0.93 | C | 0.07 | 1.01 | 0.87, 1.19 | 0.9 |
| 4 | rs115162070 | G | 0.93 | A | 0.07 | 0.90 | 0.75, 1.07 | 0.2 |
| 4 | rs11729561 | T | 0.92 | C | 0.08 | 0.96 | 0.82, 1.12 | 0.6 |
| 4 | rs112641600 | C | 0.89 | T | 0.11 | 0.83 | 0.72, 0.96 | 0.01 |
| 5 | rs62377777 | T | 0.79 | C | 0.21 | 0.96 | 0.87, 1.07 | 0.5 |
| 5 | rs4240376 | G | 0.80 | T | 0.20 | 0.98 | 0.88, 1.09 | 0.7 |
| 5 | rs10039856 | C | 0.90 | T | 0.10 | 1.10 | 0.96, 1.26 | 0.2 |
| 5 | rs2220543 | T | 0.71 | A | 0.29 | 1.03 | 0.94, 1.14 | 0.5 |
| 5 | rs113791144 | C | 0.93 | T | 0.07 | 0.97 | 0.82, 1.14 | 0.7 |
| 6 | rs6933436 | A | 0.71 | C | 0.29 | 1.01 | 0.92, 1.11 | 0.9 |
| 6 | rs10755709 | A | 0.70 | G | 0.30 | 1.11 | 1.01, 1.21 | 0.03 |
| 6 | rs140247774 | C | 0.93 | T | 0.07 | 0.93 | 0.78, 1.10 | 0.4 |
| 6 | rs16873740 | T | 0.88 | A | 0.12 | 1.16 | 1.03, 1.32 | 0.02 |
| 6 | rs9386484 | T | 0.76 | A | 0.24 | 0.95 | 0.85, 1.06 | 0.4 |
| 8 | rs118072448 | T | 0.92 | C | 0.08 | 0.82 | 0.70, 0.97 | 0.02 |
| 8 | rs10808999 | A | 0.13 | G | 0.87 | 1.01 | 0.89, 1.14 | 0.9 |
| 8 | rs13282163 | A | 0.92 | C | 0.08 | 0.93 | 0.80, 1.09 | 0.4 |
| 8 | rs11779911 | C | 0.67 | A | 0.33 | 0.99 | 0.91, 1.09 | 0.9 |
| 8 | rs2010843 | T | 0.47 | C | 0.53 | 1.04 | 0.96, 1.13 | 0.4 |
| 9 | rs3895472 | T | 0.08 | C | 0.92 | 1.03 | 0.88, 1.21 | 0.7 |
| 9 | rs12236000 | G | 0.92 | C | 0.08 | 0.95 | 0.81, 1.11 | 0.5 |
| 9 | rs7027911 | G | 0.57 | A | 0.43 | 1.10 | 1.01, 1.21 | 0.04 |
| 10 | rs71481792 | A | 0.38 | T | 0.62 | 0.89 | 0.82, 0.97 | 0.01 |
| 10 | rs2091431 | A | 0.28 | G | 0.72 | 1.04 | 0.94, 1.14 | 0.5 |
| 10 | rs1892429 | A | 0.84 | G | 0.16 | 0.98 | 0.87, 1.11 | 0.8 |
| 10 | rs10793436 | G | 0.68 | T | 0.32 | 0.94 | 0.85, 1.04 | 0.2 |
| 10 | rs1441121 | T | 0.57 | A | 0.43 | 0.95 | 0.87, 1.03 | 0.2 |
| 11 | rs10766439 | A | 0.37 | G | 0.63 | 0.97 | 0.89, 1.05 | 0.4 |
| 12 | rs11613792 | A | 0.85 | G | 0.15 | 1.01 | 0.89, 1.14 | 0.9 |
| 12 | rs12823094 | T | 0.76 | A | 0.24 | 1.08 | 0.98, 1.19 | 0.1 |
| 13 | rs1984162 | A | 0.75 | G | 0.25 | 1.10 | 1.00, 1.21 | 0.05 |
| 13 | rs12871414 | C | 0.74 | T | 0.26 | 0.95 | 0.86, 1.05 | 0.3 |
| 14 | rs2238187 | A | 0.65 | G | 0.35 | 1.07 | 0.98, 1.17 | 0.1 |
| 14 | rs12587980 | C | 0.63 | T | 0.37 | 1.03 | 0.94, 1.13 | 0.5 |
| 15 | rs12593288 | C | 0.80 | T | 0.20 | 0.91 | 0.82, 1.01 | 0.08 |
| 15 | rs2229117 | G | 0.86 | C | 0.14 | 0.90 | 0.80, 1.02 | 0.1 |
| 16 | rs72803978 | A | 0.94 | G | 0.06 | 0.88 | 0.74, 1.05 | 0.2 |
| 17 | rs34761447 | C | 0.90 | T | 0.10 | 1.02 | 0.89, 1.18 | 0.8 |
| 17 | rs178840 | G | 0.75 | A | 0.25 | 0.94 | 0.85, 1.04 | 0.2 |
| 18 | rs12958013 | T | 0.86 | C | 0.14 | 1.08 | 0.96, 1.22 | 0.2 |
| 19 | rs8105499 | C | 0.70 | A | 0.30 | 0.98 | 0.90, 1.08 | 0.7 |
| 19 | rs60744406 | A | 0.41 | G | 0.59 | 1.02 | 0.94, 1.11 | 0.7 |
| 19 | rs10411226 | G | 0.25 | A | 0.75 | 1.04 | 0.94, 1.15 | 0.5 |
| 21 | rs2252109 | A | 0.48 | T | 0.52 | 0.98 | 0.90, 1.06 | 0.6 |
| 22 | rs5757427 | T | 0.65 | A | 0.35 | 0.96 | 0.88, 1.05 | 0.4 |
| 22 | rs7290963 | G | 0.55 | T | 0.45 | 1.00 | 0.92, 1.09 | 0.9 |
| 22 | rs11090305 | T | 0.80 | C | 0.20 | 1.06 | 0.96, 1.18 | 0.2 |
| 22 | rs62220604 | G | 0.73 | A | 0.27 | 0.97 | 0.88, 1.07 | 0.5 |
| 3 | rs11385942 | G | 0.92 | GA | 0.08 | 1.16 | 1.00, 1.34 | 0.05 |
| 9 | rs657152 | C | 0.63 | A | 0.37 | 0.95 | 0.87, 1.03 | 0.2 |
| **12 SNPs from Pairo-Castineira et al. [2]** |  |  |  |  |  |  |  |  |
| 3 | rs71325088 | T | 0.92 | C | 0.08 | 1.15 | 0.99, 1.33 | 0.07 |
| 3 | rs73064425 | C | 0.92 | T | 0.08 | 1.15 | 0.99, 1.33 | 0.07 |
| 6 | rs9380142 | G | 0.30 | A | 0.70 | 1.08 | 0.99, 1.19 | 0.09 |
| 6 | rs143334143 | G | 0.93 | A | 0.07 | 1.00 | 0.85, 1.18 | 1.0 |
| 6 | rs3131294 | A | 0.13 | G | 0.87 | 1.00 | 0.88, 1.13 | 1.0 |
| 12 | rs6489867 | C | 0.36 | T | 0.64 | 0.98 | 0.90, 1.07 | 0.7 |
| 12 | rs10735079 | G | 0.36 | A | 0.64 | 0.98 | 0.90, 1.07 | 0.6 |
| 19 | rs2109069 | G | 0.68 | A | 0.32 | 1.01 | 0.92, 1.11 | 0.8 |
| 19 | rs74956615 | T | 0.95 | A | 0.05 | 1.05 | 0.87, 1.29 | 0.6 |
| 19 | rs11085727 | C | 0.72 | T | 0.28 | 1.06 | 0.96, 1.16 | 0.2 |
| 21 | rs13050728 | C | 0.68 | T | 0.32 | 1.08 | 0.99, 1.18 | 0.1 |
| 21 | rs2236757 | G | 0.70 | A | 0.30 | 1.06 | 0.97, 1.16 | 0.2 |
| **40 SNPs identified from Host Genetics Initiative [3]** |  |  |  |  |  |  |  |  |
| 1 | rs17102023 | A | 1.00 | G | 0.00 | 1.03 | 0.62, 2.74 | 0.5 |
| 1 | rs115492982 | G | 1.00 | A | 0.00 | 2.46 | 1.23, 4.91 | 0.01 |
| 1 | rs2224986 | C | 0.91 | T | 0.09 | 0.98 | 0.85, 1.14 | 0.8 |
| 1 | rs74508649 | C | 1.00 | T | 0.00 | 1.05 | 0.47, 2.34 | 0.9 |
| 1 | rs112317747 | T | 0.97 | C | 0.03 | 1.26 | 1.00, 1.58 | 0.05 |
| 2 | rs183569214 | G | 1.00 | A | 0.00 | 0.71 | 0.15, 3.41 | 0.7 |
| 2 | rs77764981 | T | 1.00 | C | 0.00 | 1.29 | 0.54, 3.10 | 0.6 |
| 2 | rs2034831 | A | 0.94 | C | 0.06 | 1.23 | 1.04, 1.45 | 0.02 |
| 3 | rs1705826 | C | 0.63 | G | 0.37 | 1.03 | 0.94, 1.12 | 0.6 |
| 3 | rs35896106 | C | 0.92 | T | 0.08 | 1.17 | 1.01, 1.35 | 0.04 |
| 3 | rs76374459 | G | 0.94 | C | 0.06 | 1.21 | 1.02, 1.42 | 0.03 |
| 3 | rs35652899 | C | 0.93 | G | 0.07 | 1.17 | 1.01, 1.36 | 0.04 |
| 3 | rs12639224 | C | 0.73 | T | 0.27 | 1.02 | 0.93, 1.12 | 0.6 |
| 3 | rs34901975 | G | 0.89 | A | 0.11 | 1.11 | 0.98, 1.27 | 0.1 |
| 3 | rs71615437 | A | 0.92 | G | 0.08 | 1.11 | 0.96, 1.29 | 0.2 |
| 3 | rs13433997 | T | 0.88 | C | 0.12 | 1.10 | 0.97, 1.24 | 0.1 |
| 3 | rs10510749 | C | 0.91 | T | 0.09 | 0.99 | 0.85, 1.15 | 0.9 |
| 3 | rs115102354 | A | 0.95 | G | 0.05 | 0.96 | 0.79, 1.16 | 0.7 |
| 3 | rs13062942 | A | 0.64 | G | 0.36 | 0.93 | 0.85, 1.02 | 0.1 |
| 3 | rs76488148 | G | 0.96 | T | 0.04 | 1.25 | 1.02, 1.52 | 0.03 |
| 5 | rs4478338 | T | 0.92 | G | 0.08 | 1.08 | 0.93, 1.25 | 0.3 |
| 5 | rs111265173 | C | 1.00 | T | 0.00 | 0.98 | 0.35, 2.67 | 1.0 |
| 6 | rs61611950 | C | 0.99 | T | 0.01 | 0.92 | 0.56, 1.51 | 0.8 |
| 7 | rs6967210 | T | 0.99 | C | 0.01 | 1.16 | 0.84, 1.59 | 0.4 |
| 8 | rs332040 | G | 0.53 | A | 0.47 | 1.00 | 0.92, 1.09 | 0.9 |
| 9 | rs71480372 | A | 0.66 | T | 0.34 | 0.99 | 0.90, 1.09 | 0.8 |
| 9 | rs74790577 | A | 1.00 | T | 0.00 | 1.05 | 0.27, 4.06 | 0.9 |
| 10 | rs5016035 | T | 0.51 | G | 0.49 | 1.00 | 0.91, 1.09 | 0.9 |
| 12 | rs7397549 | T | 0.59 | C | 0.41 | 1.00 | 0.91, 1.09 | 0.9 |
| 13 | rs2649134 | C | 0.97 | T | 0.03 | 0.93 | 0.72, 1.19 | 0.6 |
| 14 | rs144114696 | G | 1.00 | A | 0.00 | 2.49 | 0.50, 12.38 | 0.3 |
| 15 | rs77055952 | A | 0.95 | G | 0.05 | 1.07 | 0.88, 1.29 | 0.5 |
| 15 | rs74750712 | T | 1.00 | G | 0.00 | 1.33 | 0.66, 2.70 | 0.4 |
| 16 | rs72779789 | G | 0.95 | C | 0.05 | 1.03 | 0.85, 1.26 | 0.8 |
| 16 | rs145643452 | G | 0.99 | A | 0.01 | 1.03 | 0.61, 1.74 | 0.9 |
| 17 | rs9890316 | G | 0.69 | A | 0.31 | 1.01 | 0.92, 1.11 | 0.9 |
| 18 | rs142257532 | T | 0.97 | C | 0.03 | 1.01 | 0.78, 1.30 | 1.0 |
| 20 | rs56259900 | A | 1.00 | G | 0.00 | 1.11 | 0.63, 1.94 | 0.7 |
| 20 | rs76253189 | C | 0.99 | G | 0.01 | 1.01 | 0.72, 1.42 | 1.0 |
| 21 | rs75994231 | C | 0.98 | T | 0.02 | 1.06 | 0.79, 1.43 | 0.7 |

Chr, chromosome number; CI, confidence interval; OR, odds ratio per effect allele
Note: imputed data was used for the unadjusted odds ratios; the 64 SNPs used in Dite et al.[1] included rs11385942 and rs657152 from Ellinghaus et al. [4]

**Supplementary Table S2.** Characteristics of cases and controls for validation of the prototype model to predict severe COVID-19.

| Variable |  | Cases<br>N=1,234 | Controls<br>N=4,805 |
| --- | --- | --- | --- |
|  |  | Mean (SD) | Mean (SD) |
| SNP score | % risk alleles | 61.3 (3.9) | 61.4 (3.8) |
|  |  | N (%) | N (%) |
| Age group (years) | 50–59 | 187 (15.2) | 1,717 (35.7) |
|  | 60–69 | 289 (23.4) | 1,609 (33.5) |
|  | 70+ | 758 (61.4) | 1,479 (30.8) |
| Sex | Female | 529 (42.9) | 2,613 (54.4) |
|  | Male | 705 (57.1) | 2,192 (45.6) |
| Ethnicity | White | 1,137 (92.1) | 4,433 (92.3) |
|  | Other | 87 (7.1) | 359 (7.5) |
|  | Missing | 10 (0.8) | 13 (0.3) |
| ABO blood type | O | 524 (42.5) | 1,855 (38.6) |
|  | A | 537 (43.5) | 2,244 (46.7) |
|  | B | 124 (10.1) | 504 (10.5) |
|  | AB | 49 (4.0) | 202 (4.2) |
| Autoimmune disease (rheumatoid arthritis, lupus or psoriasis) | No | 1,160 (94.0) | 4,635 (96.5) |
|  | Yes | 74 (6.0) | 170 (3.5) |
| Cancer – haematological | No | 1,206 (97.7) | 4,775 (99.4) |
|  | Yes | 28 (2.3) | 30 (0.6) |
| Cancer – non-haematological | No | 978 (79.3) | 4,227 (88.0) |
|  | Yes | 256 (20.8) | 578 (12.0) |
| Diabetes | No | 965 (78.2) | 4,393 (91.4) |
|  | Yes | 269 (21.8) | 412 (8.6) |
| Hypertension | No | 577 (46.8) | 3,428 (71.3) |
|  | Yes | 657 (53.2) | 1,377 (28.7) |
| Respiratory disease (excluding asthma) | No | 937 (75.9) | 4,456 (92.7) |
|  | Yes | 297 (24.1) | 349 (7.3) |

SD, standard deviation

